# Benchtop evaluation of a double stent retriever thrombectomy technique for acute ischemic stroke treatment

**DOI:** 10.1101/2022.10.12.22280760

**Authors:** Jeremy Hofmeister, Gianmarco Bernava, Andrea Rosi, Philippe Reymond, Olivier Brina, Michel Muster, Karl-Olof Lovblad, Paolo Machi

## Abstract

**Background:** A mechanical thrombectomy technique using a double stent retriever (DSR) approach has been reported for the treatment of patients with acute ischemic stroke. The purpose of this study was to perform a benchtop evaluation of the mechanism of action and efficacy of a DSR approach compared to a single stent retriever approach.

**Methods:** *In vitro* mechanical thrombectomy procedures were performed in a vascular phantom reproducing a M1-M2 occlusion with two different clot analog consistencies (soft and hard). We compared the DSR approach to the single stent retriever approach and recorded the recanalisation rate, distal embolization, and retrieval forces of each mechanical thrombectomy procedure.

**Results:** The DSR approach achieved a higher recanalization rate and lower embolic complications compared to the single stent retriever approach. This seems to stem from two facts: the greater probability of targeting the correct artery with two stents in the case of bifurcation occlusion, and an improved clot capture mechanism using the DSR approach. However, the DSR was associated with an increased initial retrieval force.

**Conclusion:** *In vitro* evaluation of the mechanism of action of the DSR provided explanations that appear to support the high efficacy of such an approach in patient cohorts and could help operators when selecting the optimal mechanical thrombectomy strategy in cases of arterial occlusions difficult to treat with a single stent retriever.

## 1. Introduction

A mechanical thrombectomy (MTB) technique using a double stent retriever (DSR) has been recently reported as a successful approach to treat patients suffering from an acute ischemic stroke (AIS) due to the occlusion of a large vessel of the anterior circulation.[1, 2, 3, 4, 5, 6, 7, 8, 9, 10, 11, 12] Compared with a single stent retriever (SSR) technique, a DSR would ensure a larger metal surface interacting with the clot and an associated pincer effect, thus increasing the odds of successful capture. On the other hand, such an approach would lead to a greater force exerted by the two SRs over the vessel wall, potentially increasing the risks of vessel damage. In this study, we aimed to evaluate and compare the efficacy of the DSR technique with the SSR technique. In addition, we measured the forces exerted to retrieve an SSR or two SRs simultaneously in order to evaluate how such forces differed between the techniques.

## 2. Methods

### 2.1. In vitro model

We performed *in vitro* MTB procedures using a vascular phantom and clot analogs (CA) of different consistencies. A linear traction machine was used to retrieve the SRs after being deployed into the phantom over the clots. For the purpose of the study, we conceived a vascular phantom carved from a block of polymethylmethacrylate using a computer-controlled machine, which reproduced an idealized middle cerebral artery anatomy with an M1 segment branching into two M2 segments. Both M2 segments formed an angle of 120° with the M1 segment (1.A). The phantom was continuously flushed by warm water heated at 37°C using a steady state pump and the outlet of the system was equipped with a canister in order to capture potential distal emboli occurring during the experimental MTBs.

### 2.2. Clot analogs

Soft and hard CAs were produced according to the method reported by Bernava et al.[13] Briefly, CAs were produced using mixtures of guar and borax forged into cylindrical specimens and compressed by one-half of their height using a vertical compression machine (Sauter FL10, Sauter GmbH, Wutöschingen, Germany) to determine their consistencies. Samples that required a compression force of <3 mN/mm^2^ were considered as ‘soft’ and those that required a force of *>*5 mN/mm^2^ were considered as ‘hard’. After consistency measurements, CAs were modeled to have a diameter of 2.5 mm and a length of 10 mm.

### 2.3. In vitro MTB

Two operators with experience in MTB (JH and PM) performed *in vitro* MTB procedures. CAs were placed inside the vascular phantom to occlude a M1 segment and one of the two M2 branches. In each case, the portion of the CA placed in M1 completely arrested the flow of the two M2 branches. The rationale was to reproduce realistic clinical conditions where the operator is not aware of the distal location of a clot occluding M1 or M1-M2. In cases of MTB conducted with a SSR, we reproduced two experimental scenarios: one in which the SR was placed in the M2 branch that was really occluded by the clot, together with the distal portion of M1, and one in which the stent was placed in the M2 branch not occluded by the clot (Figure 1.B). Experiments were conducted via a standard 8F guide catheter (Infinity, Stryker, Portage, MI, USA) introduced into the proximal inlet of the vascular phantom. One or two SR Solitaire FR 4-20 mm (Medtronic, Irvine, CA, USA) were deployed over and beyond the CAs via a 0.021 inches microcatheter (Headway, MicroVention, Aliso Viejo, CA, USA). Subsequently, the SRs were retrieved inside the large guide catheter using a linear traction machine (Sauter THM 500N500, Sauter GmbH) and coupled with a digital dynamometer (Sauter FL10, Sauter GmbH) at a constant velocity of 0.6 mm/sec. This velocity was established by measuring the average SR withdrawal velocity recorded in preliminary tests where the traction machine was not used and where the two operators (JH and PM) simulated MTBs and manually withdrew the stent(s) outside the vascular phantom. These preliminary tests were not considered to evaluate the efficacy of the DSR, but only the velocity of CA retrieval.

**Figure 1:**
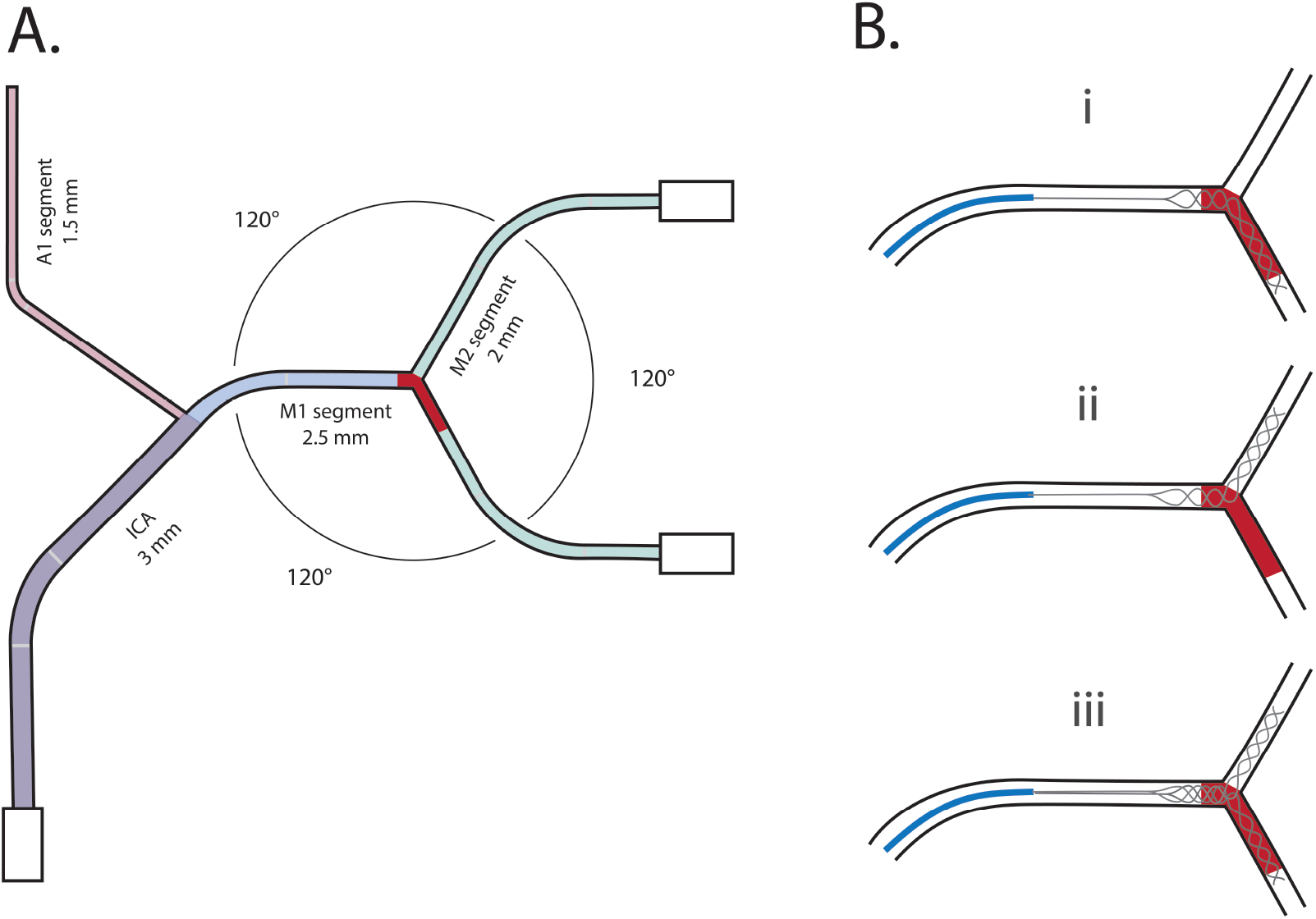
Analysis pipeline. **A**. Drawing of the vascular phantom used for the mechanical thrombectomy experiments, with vessel diameters and angles between M1 and M2 branches. **B**. Schematic representation of the experiments, with clot analog located in M1-M2 and the stent retriever placed in three different configurations positions; (i) single stent retriever placed in M1 and the M2 branch occluded by the clot; (ii) single stent retriever placed in the M1 and the M2 branch not occluded by the clot; (iii) double stent-retriever, with the stents placed in M1 and the two M2 branches.

Ten MTB procedures, including up to 3 SR passes, were performed for each of the following configurations (figure 1.B):

i. a single SR was placed from M1 to the M2 branch occluded by the CA (correct SSR);
ii. a single SR was placed from M1 to the M2 branch not occluded by the CA (wrong SSR);
iii. two SRs were placed from M1 to both M2 branches (DSR).

The recanalization rate, presence of distal emboli, and the force required to retrieve the SRs were recorded for each procedure.

### 2.4. Statistical analysis

The influences of two independent variables, *i.e*., the MTB procedure configuration (correct SSR, wrong SSR and DSR) and consistency of the CA (soft or hard) were evaluated using a two-way analysis of variance to compare their effect on the recanalization rate, distal embolization and maximal retrieval force (F*max*). Post hoc analysis using the Tukey HSD were performed in case of a significant main effect (with *≥* 2 groups) or interaction effect in order to explore differences between multiple groups on distal embolization and F*max*. All statistical comparisons were performed using R (version 4.0.5) with a significance level at 0.05 (two-tailed).

## 3. Results

### 3.1. Recanalization rate

A single SR placed from M1 to the M2 branch occluded by the CA (correct SSR) was effective in retrieving 80% of hard CAs (8/10) and 100% (10/10) of soft CAs. A single SR deployed in the branch M2 not occluded (wrong SSR) was effective in retrieving 80% of soft CA (8/10) and 0% of hard CA (0/10). The DSR technique was effective at first pass in all cases, regardless of CA consistencies (hard CA: 10/10; soft CA: 10/10). Statistical analysis showed that the recanalization rate was greater for soft than hard clots (p<0.001), and greater for DSR and correct SSR than wrong SSR (p<0.001). These analyses also showed a statistical interaction effect between CA consistencies and MTB procedure type (p<0.001), with hard CAs being easier to retrieve for correct SSR and DSR than for wrong SSR (both p<0.05).

### 3.2. Distal embolization

MTB with a single SR in the correct M2 branch did not yield distal embolization for soft CA (0/10), but resulted in 37.5% distal embolization in the case of hard TA (3/8). MTB with a single SR in the wrong M2 branch resulted in 50% distal embolization (4/8) when retrieving soft CAs. No case of distal embolization was recorded for DSR technique, regardless of CA consistencies (hard TA: 0/10; soft TA: 0/10). Statistical analyses showed an interaction effect between CA consistencies and the SR-MTB procedure for distal embolization (p<0.05), with more distal embolization in the correct SSR than in the DSR for hard CA (p<0.05), and more embolization for the wrong SSR than both correct SSR and DSR for soft CA (both p<0.05).

### 3.3. Retrieval forces

The force required to retrieve the SR varied according to the procedure configuration (p<0.001), with higher forces recorded for the DSR technique compared to a SSR in both the correct (p<0.05) and wrong branches (p<0.05) (figures 2 and 3). In the case of the DSR technique, the higher force (F*max*) was recorded at the beginning of the withdrawal when the distal portions of the SRs were located in the M2 branches. Once both SRs entirely reached M1, the force considerably decreased (figure 3). CA consistency influenced the force needed to retrieve the SRs according to the MTB configuration (p<0.05). For soft CAs, the forces were significantly higher when performing the DSR technique compared to a SSR in the correct M2 branch (p<0.05). In addition, forces were higher to retrieve soft CA by the SSR technique in the wrong M2 branch compared to SSR in the correct M2 branch (p<0.05). For hard CA, forces were significantly higher when performing DSR than both SSR in the correct (p<0.05) and the wrong branches (p<0.05).

**Figure 2:**
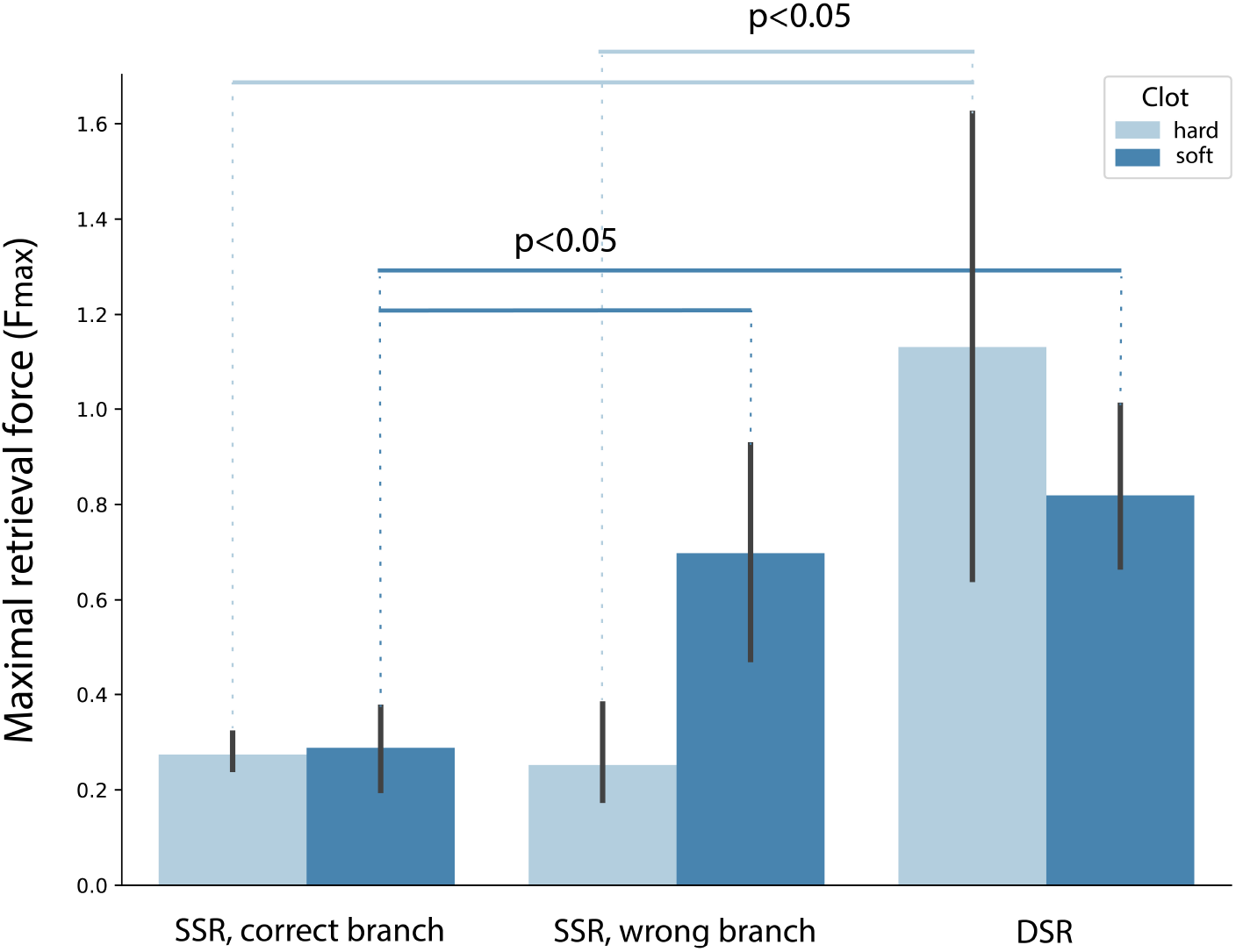
Maximal Retrieval Forces. Maximal retrieval force (in N) during the stent retriever-based mechanical thrombectomy procedures, depending on procedure type and CA consistencies. Solid lines represent statistically significant comparisons (p<0.05).

**Figure 3:**
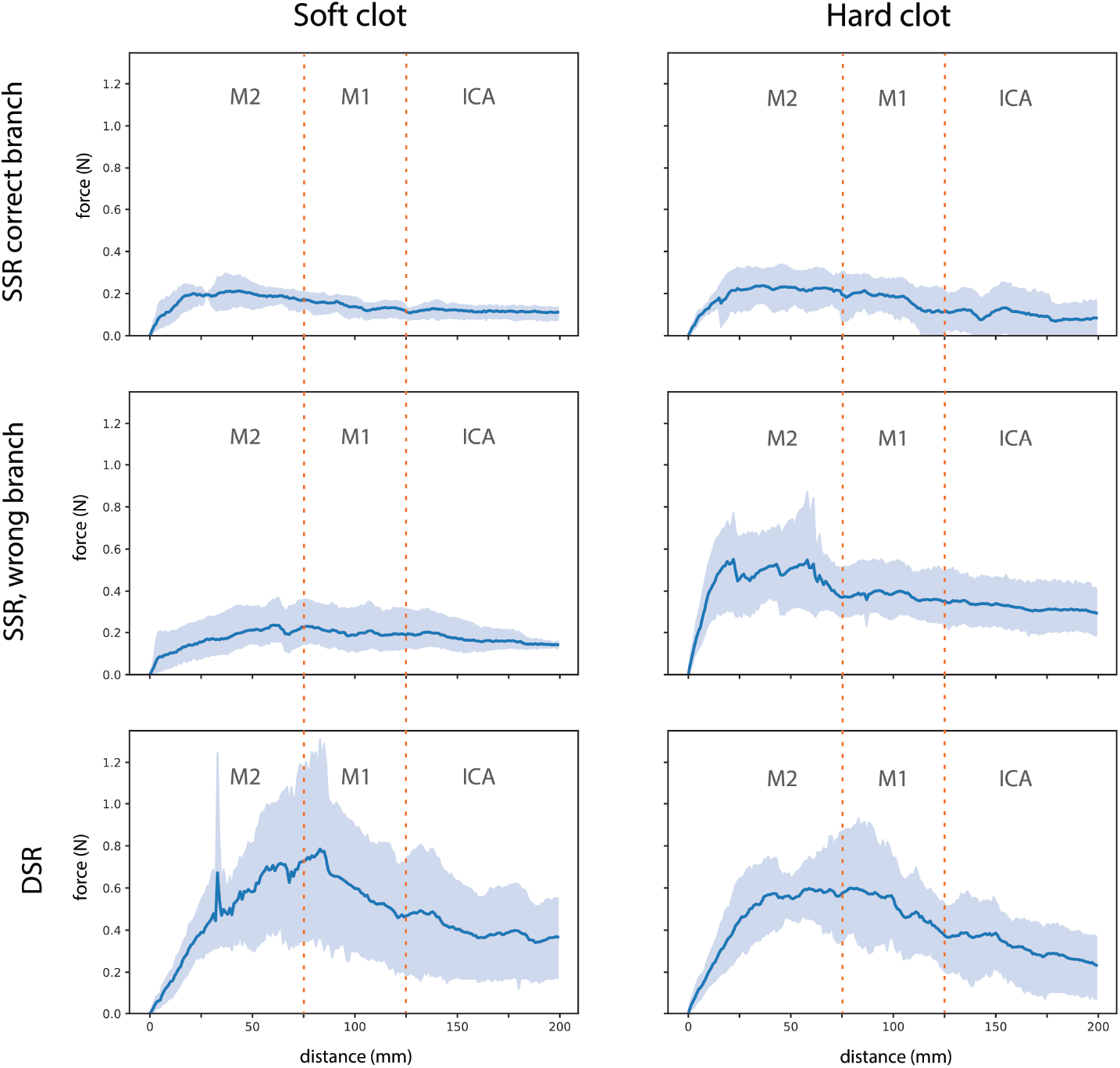
Retrieval Forces. Retrieval force (± 95% confidence interval in light blue) during the whole stent retriever-based mechanical thrombectomy procedures, depending on procedure type (DSR, correct SSR or wrong SSR) and CA consistencies (soft versus hard).

**Table 1:**
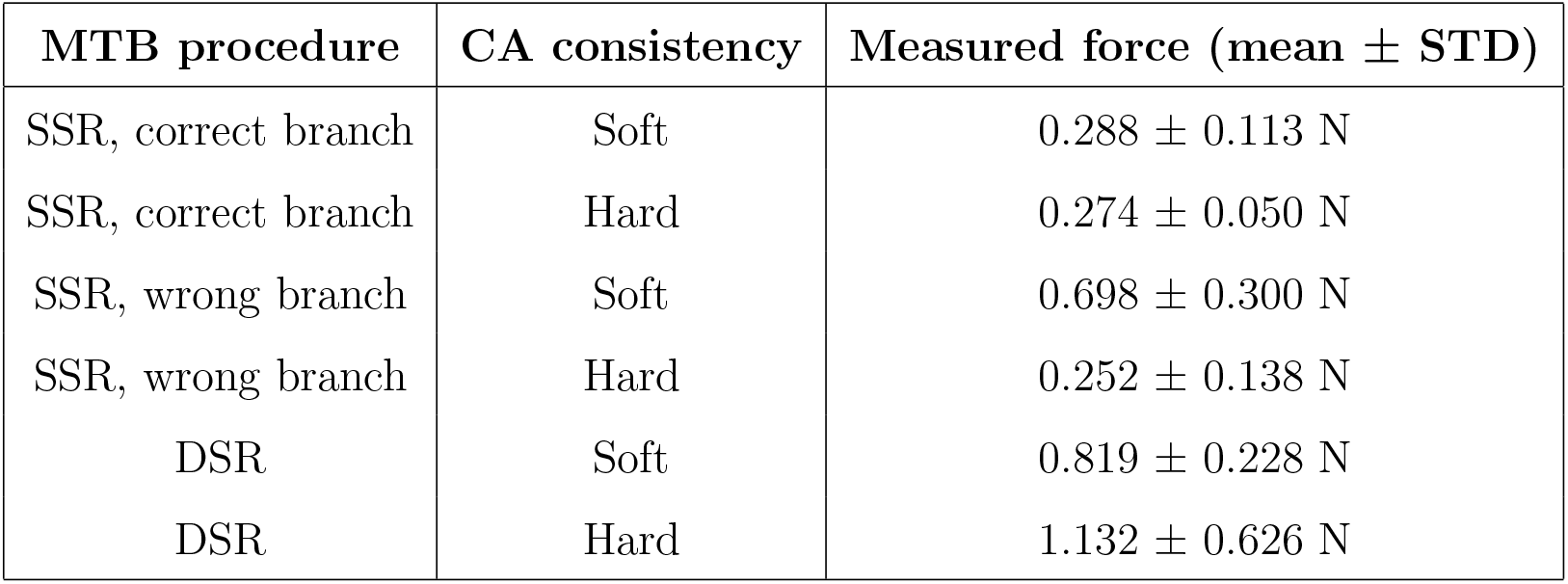
Maximal retrieval force during *in vitro* mechanical thrombectomy.

| MTB procedure | CA consistency | Measured force (mean ± STD) |
| --- | --- | --- |
| SSR, correct branch | Soft | $0.288 \pm 0.113$ N |
| SSR, correct branch | Hard | $0.274 \pm 0.050$ N |
| SSR, wrong branch | Soft | $0.698 \pm 0.300$ N |
| SSR, wrong branch | Hard | $0.252 \pm 0.138$ N |
| DSR | Soft | $0.819 \pm 0.228$ N |
| DSR | Hard | $1.132 \pm 0.626$ N |

## 4. Discussion

Our *in vitro* experiment allowed to evaluate the mechanism of action and efficacy of a DSR-based MTB technique. We compared the DSR approach to a conventional SSR approach in an experimental model of a simple sylvian bifurcation in order to avoid biases related to the angles of the M1-M2 bifurcation. We observed that the DSR appears to be highly effective in removing clots independent of their consistency and reduces the risk of embolic complications compared with an SSR approach.

Several explanations seem to support these observations. The use of the DSR reduces the risk of targeting the wrong M2 branch. Sylvian bifurcation occlusions obscure M2 branches because of a localized clot in the distal part of the M1 segment, thereby blinding the operator to the M2 branch containing the clot and resulting in the inability to properly deploy an SSR. We observed that when an SSR is deployed in the wrong M2 branch, it does not remove any hard clot. In our *in vitro* model, we observed a good soft clot removal rate with an SSR deployed in the wrong M2 branch, but a significant risk of clot fragmentation, leading to embolic complications. Compared with the SSR, DSR increases the likelihood of targeting the M2 branch containing the clot and performing an effective MTB manoeuvre. Indeed, we observed first-pass recanalization in each MTB trial, regardless of clot consistency and without any embolic complications. Interestingly, the SSR deployed in the correct M2 branch containing the clot had a good recanalization rate for soft clots, but a slightly lower rate in the case of hard clots compared with a DSR.

Furthermore, the clot capture mechanism by DSR appears to be different from that of a SSR due to a “pincer effect” capturing the clot between the two SRs and a wider “fishing net” ensuring a low risk of clot fragmentation during the retrieval. Accordingly, we observed that the DSR decreases the risk of embolic complication compared with the SSR, even when the SSR is deployed in the correct M2 branch in the case of hard clot. Indeed, we observed a significant number of embolic complications when using an SSR for soft clots when deployed in the wrong M2 branch and for hard clots when deployed in the correct M2 branch. Whereas SSRs integrate into soft clots and drive hard clots into a rolling phenomenon, the addition of a second SR changes the clot capture mechanism in two ways (wider “fishing net” and “pincer effect”), regardless of clot consistency.

One question that arises with the use of the DSR is its danger to the arterial wall, which could increase the risk of hemorrhagic complications compared with an SSR. Although our study was not designed to answer this question definitively, we measured the retrieval forces of the SRs during all MTB manoeuvres to estimate whether these forces were excessively higher with a DSR compared with an SSR, and to gain insight into the interaction of the devices with the arterial walls during MTB. We observed that the DSR approximately doubles the retrieval force compared with an SSR when the SRs are in the M2 segments. However, this withdrawal force decreases rapidly when the SRs arrive entirely in M1. The withdrawal forces observed for an SSR are similar to those reported *in vitro* by other authors who also tested the Solitaire (4×20 mm) for MTBs in M1.[14] When using a DSR with two Solitaires (4×20 mm), we observed a higher withdrawal force than with a single Solitaire (4×20 mm), but this force was close to that measured with SSR using other devices, such as the pREset (4×20 mm). This suggests that the choice of devices for the DSR likely has an impact on the retrieval forces and safety of this approach. A multicenter analysis of DSR clinical cases has indeed shown that the choice of SR size influences the effectiveness of MTB, but that large diameter SRs lead to more hemorrhagic complications.[2] However, further studies are needed to evaluate the precise effect of the DSR on arterial walls during MTB, e.g., using histological analyses in animal models as discussed by Nogueira et al.[15]

The DSR approach has been reported through case reports of patients treated for refractory anterior circulation occlusions[1, 3, 4, 8, 11] and in rare cases of posterior circulation occlusion.[10] DSR was first reported as a rescue technique in three consecutive, retrospective patient cohorts, two single center trials comprising 28 and 10 patients, respectively[6, 5] and one multicenter study of 20 patients.[2] These studies reported final recanalization rates between 80% and 85.7% (mTICI *≥*2b/3), respectively, and a recanalization rate of 70% at the first DSR pass for the multicenter study. These recanalization rates are high when considering that these were clots that were not removed by MTB with SSR. The DSR was also recently reported as a front-line technique in anterior circulation occlusions in a single-center cohort of 39 patients, with a first-pass effect of 69% and a final recanalization rate of 100%.[12] The high rates of full recanalization by DSR in these clinical cohorts are close to the results we observed *in vitro*. However, the fact that all clots are retrieved by DSR at first pass *in vitro* is superior to what is observed in patients, where several passes may be required. This might relate to the inherent differences between *in vivo* and *in vitro* studies. Indeed, in our experiment, we used a simplified model of middle cerebral artery bifurcation where two M2 segments formed the same angle (120°) with the M1 segment to ensure the same interaction between the clot and the SR in each M2 branch. Such a favorable interaction probably does not always occur *in vivo* because of the variability of MCA bifurcations in humans.[16] The retrospective cohorts mentioned above reported symptomatic hemorrhagic complication rates between 7.9% and 10%. These rates are higher than those reported in the recent literature for SSR approach and may be related to the presence of a larger surface area of metal interacting with the vessel wall, which could result in the increased initial retrieval force that we observed with DSR.

## Data Availability

All data produced in the present study are available upon reasonable request to the authors.

## 5. Conclusion

Our *in vitro* study evaluating the mechanism of action of DSR appears to provide explanations that support results observed in previous patient cohorts. The high recanalization rate and low risk of embolic complications observed with DSR seem to stem from the greater probability of targeting the correct artery in the case of bifurcation occlusion, as well as an improved capture mechanism.

